# Human genotype-to-phenotype predictions: boosting accuracy with nonlinear models

**DOI:** 10.1101/2021.06.30.21259753

**Authors:** Aleksandr Medvedev, Satyarth Mishra Sharma, Evgenii Tsatsorin, Elena Nabieva, Dmitry Yarotsky

## Abstract

Genotype-to-phenotype prediction is a central problem of human genetics. In recent years, it has become possible to construct complex predictive models for phenotypes, thanks to the availability of large genome data sets as well as efficient and scalable machine learning tools. In this paper, we make a three-fold contribution to this problem. First, we ask if state-of-the-art nonlinear predictive models, such as boosted decision trees, can be more efficient for phenotype prediction than conventional linear models. We find that this is indeed the case if model features include a sufficiently rich set of covariates, but probably not otherwise. Second, we ask if the conventional selection of single nucleotide polymorphisms (SNPs) by genome wide association studies (GWAS) can be replaced by a more efficient procedure, taking into account information in previously selected SNPs. We propose such a procedure, based on a sequential feature importance estimation with decision trees, and show that this approach indeed produced informative SNP sets that are much more compact than when selected with GWAS. Finally, we show that the highest prediction accuracy can ultimately be achieved by ensembling individual linear and nonlinear models. To the best of our knowledge, for some of the phenotypes that we consider (asthma, hypothyroidism), our results are a new state-of-the-art.

## 1 Introduction

The problem of predicting phenotype from genotype is a “holy grail” of modern genetics, with practical applications in fields such as personalized medicine [1] and genomic selection for agriculture [2], and is an active area of research. Its relevance has grown with the affordability of genotyping, and will likely continue to increase as sequencing becomes more commonplace.

Classical predictive models for human phenotypes from full genotypes are variations of linear methods; a popular approach is to use a method that combines regression with suitable regularization (e.g., Lasso [3]). Due to the size of the data and computational limitations, regression is typically preceded by feature selection through genome-wide association study (GWAS) methods which identify the features (Single Nucleotide Polymorphisms, SNPs) most significantly associated with the phenotype, although methods that do not always need this step are being developed [4, 5]. Linear models work well with the large number of SNPs, many of which have small effects, and with the limited amount of data (available genotype-phenotype pairs). In a striking success of linear methods in the genotype-to-phenotype prediction problem, polygenic risk scores that are computed on the basis of a person’s entire genotype, have been shown to be as well or more sensitive in identifying individuals susceptible to certain diseases as Mendelian risk genes [6].

At the same time, non-linear effects, such as epistasis, can be a significant factor contributing to a phenotype [7]; consequently, there are indications that models which account for non-linearity, for examples for interactions between features, can have better prediction accuracy and may alleviate the problem of lack of the generalizability of genotype-based predictions across genetic backgrounds, at least on simulated data and/or model organisms [8, 9].

If nonlinear effects do indeed contribute significantly to some phenotypes, then latest-generation machine learning methods, which take nonlinearity into account, should outperform linear ones. These methods require large amounts of training data, which is now becoming available through datasets encompassing hundreds of thousands of individuals, such as the UK Biobank [10]. So far, the application of nonlinear machine-learning methods to the phenotype prediction problem has been inconclusive [11–13]. For some phenotypes, the effects may indeed be overwhelmingly additive and nonlinear methods may not contribute much (height is believed to be one such example [14]), while others may have a genetic architecture involving interactions both among genotypes and between genotypes and other covariates such as age.

This discussion motivates three questions that we address in this paper.

### Nonlinearity

Is there any nonlinearity in the human genotype-phenotype relationship that can be efficiently exploited by state-of-the-art machine learning methods to reliably improve prediction accuracy?

### SNP selection

How optimal is the state-of-the-art pipeline of GWAS-based SNP selection followed by a predictive model? Can it be improved by incorporating the nonlinearity into the selection step of the prediction pipeline?

### Most accurate models

What are, ultimately, the most accurate models for human genotype-to-phenotype prediction? How much of an improvement over the current state of the art can we achieve by using nonlinear methods, adjusting the pipeline, and aggregating results of different methods?

## 2 Our contribution

We address the above questions by considering several commonly studied human phenotypes and systematically exploring how the prediction accuracy is affected by the model type (linear or gradient boosted decision trees), the strategy of feature selection, and the strategy of aggregating (ensembling) optimal individual models.

### Nonlinearity

Our main tool in examining nonlinearity is experiments with gradient boosted decision trees of different depth, as implemented by the library XGBoost [15]. Trees of depth 1 depend linearly on SNPs, while deeper trees depend on them nonlinearly, which allows us to directly compare models with or without nonlinearity (see Section 3.2). In addition, we compare these results with the performance of the state-of-the-art linear method, Snpnet [4]. Moreover, we analyze the significance of nonlinear effects with respect to the number of additional (non-SNP) covariates included in the model, and with respect to interactions between features within the same group or between different groups (SNPs, covariates).

### SNP selection

We propose a new approach to SNP selection, based on constructing a preliminary lightweight XGBoost model (see Section 3.4). Our approach selects different set of SNPs which is 2-5 times smaller than GWAS but achieves a similar prediction accuracy.

### Most accurate models

We identify the top performing models constructed by individual algorithms such as XGBoost and Snpnet, and combine them to further improve the prediction accuracy. As usual in machine learning, the most accurate models are obtained by ensembling a number of (preferably, sufficiently diverse) more simple models. We consider several ensembling strategies (see Section 3.3).

## 3 Methods

### 3.1 Lasso and Snpnet

Most current methods for the prediction of phenotype from genotype are based on some form of penalized or Bayesian regression [16]. Lasso [3], which is linear regression with an 𝓁_1_-norm penalty, is well-suited for this task as genotype matrices are compressed sensors [17] and are sparse with respect to almost any phenotype. Lasso uses an absolute value regularization of parameters and tends to drive most of them to zero, thus selecting only a relatively small number of features.

Snpnet is a modification of Lasso that enables finding exact solutions to extremely high dimensional multivariate regression tasks on large datasets through an iterative batched screening process [4]. As Snpnet achieves state of the art performance in the prediction of several phenotypes, we opted to choose it as a baseline linear method to compare the performance of nonlinear methods to.

### 3.2 XGBoost

XGBoost [15] is a well-known implementation of gradient boosted decision trees [18]. XGBoost iteratively adds trees to the model by optimizing the following objective at each step *t*:

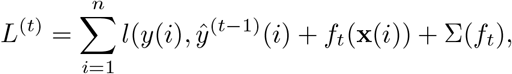

where **x**(*i*) is the input feature vector for the instance *i*, 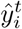 is the prediction for the instance *i* at step *t, l* is the loss function, *f*_*t*_ is the new decision tree, *f*_*t*_(**x**_*i*_) are the new tree predictions and Σ(*f*_*t*_) is a penalization term for the complexity of the new tree. The full XGBoost model is the sum of all the constructed decision trees. Each split in the tree is performed by comparing one of the features to a threshold value. We can interpret a learned XGBoost model by inspecting the trees it consists of and the features they split on, with each feature being a particular SNP or covariate. An example of such a tree with depth 2 is shown in Figure 1. Values in leaf nodes represent the influence of these particular combination of SNPs and their values on the phenotype.

**Fig 1.**
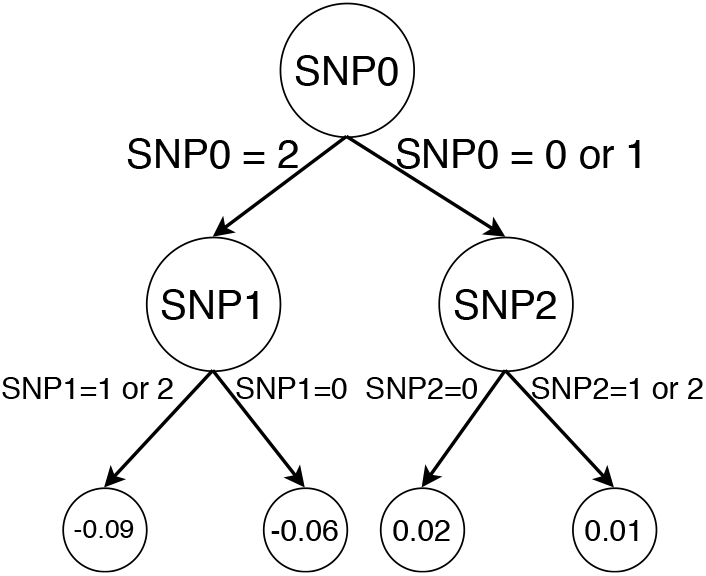
Example of a depth-2 decision tree that uses three different SNPs. Leaf nodes represent the weights that these particular SNP values contribute to the value of final prediction for a single datapoint. The genotypes are encoded as 0: homozygous reference, 1: heterozygote, 2: homozygous alternative.

It is important to note that an XGBoost model of depth 1 is a sum of *univariate* functions (i.e., depending on a single input feature *x*_*k*_, e.g., one SNP). Conversely, any sum of univariate functions can be represented (or approximated, in the case of continuous variables) by a depth-1 XGBoost model. (This is so because any univariate function *f* (*x*) can be represented or approximated by linear combinations of the indicator functions **1** {*x > c*} with various thresholds *c*.) In this sense, depth-1 XGBoost models are equivalent to *linear* models defined on one-hot-encoded input variables. For example, if different features 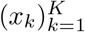 represent *K* SNPs with values 0,1,2, then a general depth-1 XGBoost model on these features is equivalent to the linear model

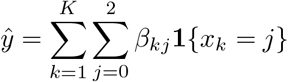

with arbitrary coefficients *β*_*kj*_.

In contrast, XGBoost models of depth 2 or higher are able to consider pairwise interactions between different features. It is generally expected that a part of phenotype variance is due to epistatic interactions between SNPs [19]; also, there might be interactions involving environmental features (covariates) used in the prediction. Accordingly, we can test how much predictions benefit from including pairwise interactions by comparing depth-1 and higher-depth XGBoost models. In this work we consider only depths 1 and 2, because larger depths do not bring noticeable improvement on our data.

XGBoost can naturally incorporate some pairwise epistatic interactions of SNPs by placing them in one tree. For example, in Figure 1 it can assign any effect sizes to the combinations of (SNP0 = 2 and SNP1 = 1 or 2) and (SNP0 = 2 and SNP1 = 0). If there is an epistasis in the form *y* = *β*_*ep*_ · *I*_*SNP* 0=2_ · *I*_*SNP* 1=0_ (where *I* denotes an indicator variable), then the XGBoost tree in Figure 1 can catch it. In this case, *β*_*ep*_ is the effect of interaction between SNP0=2 and SNP1=0 on phenotype *y*.

A useful feature of XGBoost is its ability to restrict interaction between sets of features in single trees and thus enable the study of feature interactions. By allowing or disallowing XGBoost to combine genotype and environment features in the same trees, we can test the effects of single SNP–single environment feature interactions while controlling for model expressivity in other respects.

Another useful feature of XGBoost is the reporting of feature importance scores, which is used in the XGBoost-based SNP selection approach that we propose in this paper.

An important advantage of XGBoost over more complex methods such as neural networks is that it has only a small number of important hyperparameters and is accordingly easy to tune. One of these hyperparameters is the tree depth; as already noted, we set it to 1 or 2. Another important hyperparameter is the number of trees; in our experiments we monitor the XGBoost performance throughout learning and select the optimal number of trees. We observe the other hyperparameters to provide the optimal performance at their default values, except the *l*_1_ regularization parameter *α* as in Lasso. We use *α* from 15 to 20 for all our models.

### 3.3 Ensembling and stacking

We can combine predictions obtained from several different models in order to obtain a more accurate predictor that may mitigate the shortcomings of each individual model. This is a commonly used technique in machine learning called ensembling [20]. We investigated how constructing an ensemble of predictions from different models can be used to improve the overall prediction accuracy.

We considered several ensembling strategies starting with a simple ensemble with unweighted averaging of the predictions of the individual models, which shows a modest but consistent improvement in performance. This strategy proves to be less effective when the predictive models show significantly different accuracy values. In this latter case we employ weighted averaging of predictions, with weights estimated by maximizing the accuracy of the ensemble on the validation set. This approach can be viewed as a basic example of “stacking” [21] (i.e., a collection of initial models is “stacked” with a subsequently learned linear model).

One can consider more complex stacking strategies, aggregating predictions of initial models in a nonlinear way while also possibly taking additional covariates into account. In our experiments, however, these complex stacking strategies did not outperform the simpler models, and notably added additional complexity in terms of hyperparameter selection. For this reason, we limited ourselves to the simple unweighted ensemble and the basic linear stacking as described above.

### 3.4 SNP selection by XGBoost

Addressing the question of optimizing the data processing pipeline, in this work we propose a new SNP selection method based on XGBoost as an alternative to GWAS-based SNP selection. A typical human genotype dataset has at least 600K called SNPs, and in many cases up to 90 million imputed SNPs. Moreover, UK Biobank recently released 200K exomes with more than 10 million SNPs [22].

The problem is that machine learning prediction models are not suited for handling millions of features without some feature selection. For example, phenotype prediction models typically use up to 10-100K SNPs, and these SNPs are preselected by GWAS. The largest phenotype prediction model available is Snpnet [4], which is able to use roughly 650K SNPs as features for the full UK Biobank dataset.

A drawback of GWAS-based SNP selection, however, is that the SNPs are selected based solely on their individual p-values, regardless of how much information an SNP brings compared to the other SNPs. As a result, we can expect SNPs strongly correlated with the predicted phenotype to be overrepresented in the selected subset, and the weakly correlated SNPs to be underrepresented. This is exactly the issue that we address.

Our XGBoost-based pipeline selects a balanced, compact and mutually uncorrelated subset of relevant SNPs from the whole available dataset. For that, we split the sequence of all SNPs into multiple disjoint windows. Then, we fit a separate XGBoost prediction model on each window moving either from chromosome 1 to X or from X to 1. In each window, we calculate XGBoost feature importance scores and use them for the final SNP selection. Also, every new XGBoost window model uses predictions from the previous window as the starting point: it helps the model to select new SNPs uncorrelated with SNPs selected earlier. See Figure 2 for an illustration. In the case of UK Biobank data with the total of 700K SNPs, we use windows of size 10K.

**Fig 2.**
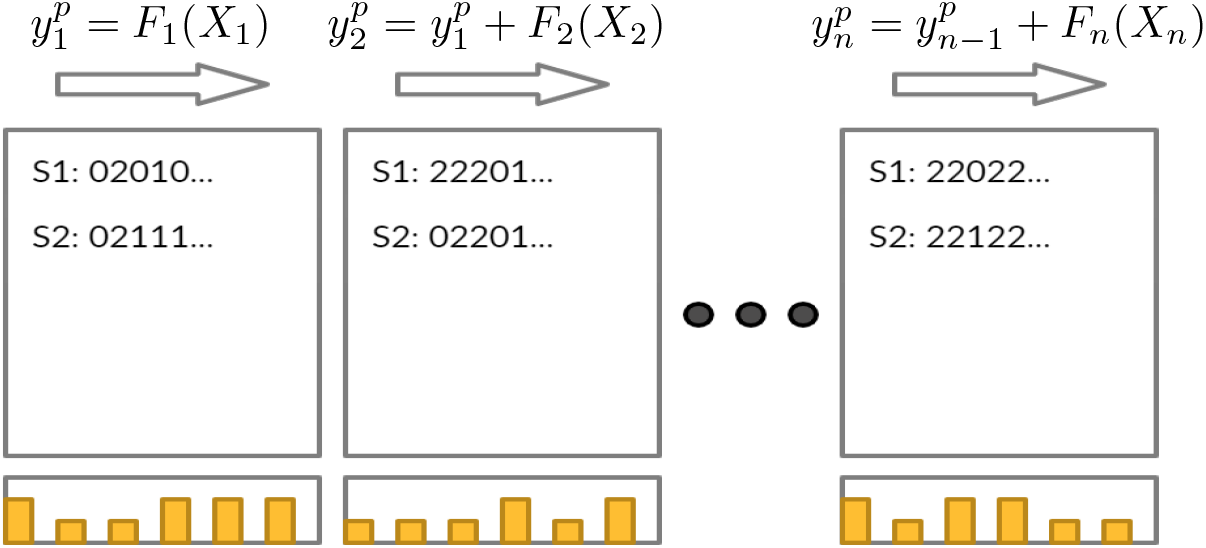
The iterative scheme of our XGBoost selection algorithm. Yellow bars are individual SNP importances. S1, S2 are individual samples. *F*_*n*_ is the *n*’th XGBoost model, using the respective window of features.

#### 3.4.1 Simulated phenotypes

To assess the performance of our XGBoost selection (namely, its ability to detect epistatic SNPs, to catch SNP-environment interactions, and to work with the nonlinearity of the environment), we performed a number of experiments with simulated phenotypes, in addition to the main set of experiments with actual phenotypes. A simulated phenotype is modelled as a weighted linear combination of linear effects **y**_*l*_, pairwise epistatic effects **y**_*ep*_ and random noise **y**_*ϵ*_:

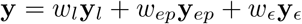

Here, **y, y**_*l*_, **y**_*ep*_, **y**_*ϵ*_ are sample-dependent, while *w*_*l*_, *w*_*ep*_, *w*_*ϵ*_ are sample-independent weights that sum to 1 and are used to adjust the relative contribution of different terms. The linear effects **y**_*l*_ linearly depend on individual SNPs, while the epistatic effects **y**_*ep*_ are formed by events involving random pairs of SNPs. See Section S2 for simulation details.

### 3.5 Metrics and error estimation

We predict several numerical as well as categorical (binary) phenotypes, and assess their accuracy using the standard *r*^2^ and ROC AUC metrics, respectively. Since our models have accuracy close to the state-of-the-art, it is important to carefully estimate the standard errors of these metrics. We use two approaches for that: one based on subsampling and another on the explicit form of the ROC AUC statistic (for categorical phenotypes). The two approaches produce close values of standard errors. See Section S1 for details.

### 3.6 Performance

The XGBoost package has GPU support and works well with missing data [15]. We built XGBoost models using one GPU node on a cluster with Nvidia Tesla V100 16GB and up to 200GB RAM [23]. In our experiments, XGBoost on this GPU was 3-6 times faster than on a CPU with 24 CPU cores. Training a single XGBoost model on GPU with 10K SNPs and 350K samples requires roughly 15GB of GPU memory and up to 80GB of RAM. The XGBoost selection step takes 1.5 hours, while building the final prediction model takes 2-3 hours. In comparison, GWAS takes 10-15 minutes on 24 CPU cores, and Snpnet requires up to 1.5 hours on 24 CPU cores on the same data.

## 4 Data

### 4.1 Dataset and Preprocessing

The genotype and phenotype data for our experiments were obtained from the full release of the UK Biobank [10]. We selected a cohort of 429,351 individuals of white British ethnicity according to data-field 21000 ‘Ethnic background’ of the biobank so as to maintain a homogeneous population structure as in [4, 5, 24, 25]. This cohort was further subdivided into train, validation, and test sets. Our train set has 343,481 samples, validation and test sets have 42,935 samples each. A total of 701,347 variants remained after filtration for allele frequency with a cutoff of 0.5%. We opted not to filter variants by linkage disequilibrium as our methods can handle its presence. For each phenotype, variant selection by GWAS was performed using PLINK 2.0 [26] taking into account age, sex, and 10 genotypic principal components. Principal components were calculated with FlashPCA [27] using the training dataset exclusively. For PCA calculation we filtered SNPs by linkage disequilibrium as recommended by the FlashPCA authors. We did not use the principal components provided by the UK Biobank because those components are based on the entire dataset and therefore would violate the training vs validation vs hold-out dataset split.

#### 4.1.1 Phenotypes

We selected two continuous and three binary phenotypes for our analysis: height, eBMD (heel-bone mineral density), asthma, hypothyroidism and psoriasis. Height is known to be highly heritable [28], and it was predicted with high accuracy by linear models in [4, 29]. eBMD also has a relatively high heritability [30, 31] and there are efforts to predict it in [24] and [32] with up to *r*^2^ = 0.25 and *r*^2^ = 0.12. Asthma and hypothyroidism have a high prevalence (11% and 5% in the UK Biobank data) and are predicted in [4, 25]. Psoriasis was selected to check how our methods work with a highly imbalanced phenotype (1% prevalence).

Only eBMD was preprocessed before prediction, using the same procedure as in [24, 32]. Additional 27 features for the eBMD prediction were selected using osteoporosis risk factors, because osteoporosis is defined as low eBMD value. These risk factors include age, sex, physical activity and alcohol consumption data, with the full list provided in S4. Asthma, psoriasis and hypothyroidism were taken from the “Non-cancer illness code, self-reported” field 20002 of the UK Biobank main dataset. For each of three categorical phenotypes, we selected age, sex and 18 additional features. The resulting set of features is different for different phenotype. We did not select any additional features besides age and sex for height, because height is predicted very well from genotype data. Also, for each phenotype we added the same 10 leading principal components calculated using FlashPCA on train dataset.

## 5 Results

### 5.1 Individual models and phenotypes

We built three groups of models for each phenotype. The first group of models uses only genetic data and sex as a covariate (because sex is also a genetic feature). The second group uses sex, age and top 10 principal components from the genotype matrix built on the training set. The third group is our attempt to make the best possible prediction and find out some nonlinear dependencies between genotype and environment, and environment and phenotype. Models of this group use age, sex and 19 additional covariates (for asthma, psoriasis and hypothyroidism) or 27 covariates (for eBMD). We chose not to create the third group for height because it is too hard to choose a set of covariates without strong two-way interaction.

Each group includes four “individual” models, and the last group additionally includes two ensemble models built using the individual models of this group. Out of the four individual models, the first two are XGBoost models of depth 1 and 2. They are trained on the set of SNPs selected by XGBoost as described in Section 3.4. The other two individual models are linear Lasso models built by Snpnet on the set of GWAS–detected SNPs. One of them uses the same number of SNPs as the XGBoost models to demonstrate that XGBoost selection–based models need fewer SNPs than GWAS-based SNPs. The last Snpnet model is trained without limiting the number of SNPs. However, while Snpnet is quite scalable, we still found it difficult to train it on more than 50K SNPs on our hardware, so the number of SNPs for this model is 50K (in most cases) or 20K (if the 50K model does not display any AUC or *r*^2^ improvement over 20K). Figure 3 shows an overview of our results. See Tables S2 and S3 for numerical values.

**Fig 3.**
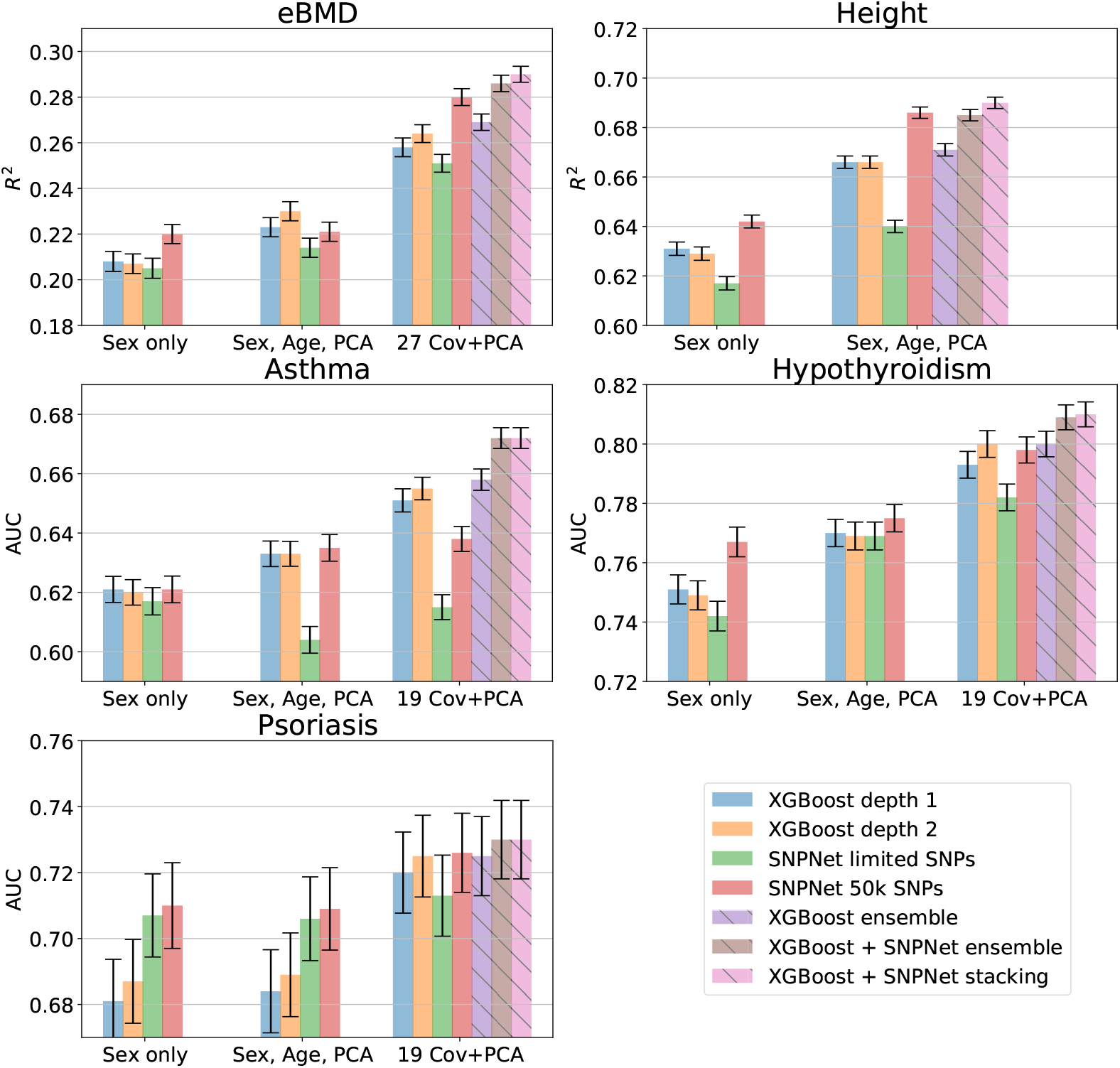
Prediction performance for all five phenotypes. Performance metrics (*r*^2^, AUC) and their standard deviations are computed on an independent test set. The total height of error bars is two standard deviations.

#### 5.1.1 Asthma

For asthma, results of the four individual models were similar in each of the three groups. We observe almost no difference between XGBoost depth 1 and depth 2, meaning that the influence of epistatic interactions is very small or non-existent in SNPs selected by XGBoost. The results in the second group show a similar picture. However, this is not the case for the last group, where XGBoost significantly outperforms GWAS-based Snpnet. Constructing ensemble of two XGBoost depth 2 models based on forward and backward SNP selection and one Snpnet model yields the best result for asthma: AUC 0.67. It demonstrates the utility of XGBoost models for asthma but Snpnet still helps us to get a better ensemble. This result is the best known to us for asthma, although every other attempt of predicting asthma in literature uses their own scheme of splitting and filtering the data. For example, Qian *et al*. [4] *obtained an AUC of 0*.613 using Snpnet, however they did not use any covariates other than age, sex, and principal components of the genotype matrix. Lello *et al*. [25] *report an AUC of 0*.632, but on genetic data only and on a test set composed of self-reported white but non-genetically British individuals from the UK Biobank.

#### 5.1.2 Hypothyroidism

Results for Hypothyroidism are generally different from asthma. For genetic data only, Snpnet outperforms both XGBoost models. Adding age and 10 principal components allows XGBoost model to catch up with Snpnet. XGBoost starts to outperform Snpnet with a limited number of SNPs only when adding 19 more covariates. The ensemble is still the best model and has an AUC of 0.807, which is also the best result known to us up to this date. Lello *et al*. [25] *report an AUC of 0*.705, however the experimental set-up differs as they use imputed genotype data only to train their model. The discussion of different splits and filtering criteria is still applicable here. Another interesting moment is that XGBoost of depth 2 outperforms XGBoost depth 1 model with full set of covariates. That could mean that there are some nonlinearities in genotype-environment or environment-phenotype interactions.

#### 5.1.3 Psoriasis

Psoriasis is a highly unbalanced phenotype with prevalence roughly 1.1%. For such an unbalanced phenotype AUC can be high even for a classifier with low precision. Also, a large change in the number of false positives can lead to a small change in the false positive rate used in ROC analysis. That’s why our AUC std estimation gives a rather high value in this case, see S1. Snpnet is still better than XGBoost for small number of covariates, but generally these results are inconclusive. Ensembles are slightly better than individual models, but the improvement is insignificant (0.1 − 0.2 standard deviations).

#### 5.1.4 eBMD

For eBMD, the unlimited version of Snpnet is generally better than XGBoost in all three groups. XGBoost of depth 2 is better than depth 1 for data with covariates. This also suggests some nonlinearity related to environment. Although Snpnet works significantly better for 27 covariates, adding XGBoost models to the ensemble further improves the result, to *r*^2^ = 0.286. This result is also state-of-the-art, to the best of our knowledge. The best eBMD prediction is in [24], *r*^2^ = 0.246.

#### 5.1.5 Height

Height is the well-known almost linear phenotype which is easy to predict. Our results generally agree with this expectation. XGBoost of depth 1 is even a bit better for genotype-only data. On the height data, XGBoost selection demonstrates its power to select a smaller set of SNPs (10K) which gives the same prediction power as bigger GWAS sets. Ensembles are still the best. That means that XGBoost and Snpnet recover slightly different prediction models.

Our result of 0.676 for the Snpnet is slightly worse than the one reported in the Snpnet paper [4]. This can be attributed to the fact that they used 650K genotyped SNPs without any GWAS preselection step, and that height is a highly additive phenotype. Also, the authors of Snpnet used a slightly different procedure of data splitting and filtering.

### 5.2 Ensembles

We constructed ensembles using three individual models for each phenotype: the best performing Snpnet model on GWAS selected SNPs, and two separate XGBoost models trained on SNPs selected by XGBoost during forward and backward passes along the genome. These ensembles generate a final prediction based on a weighted sum of the predictions of each individual model. Figure 4 shows a simplex plot in which we see how the test metric changes with varying weights assigned to each model for asthma (the weights of ensembled models correspond to the barycentric coordinates of a point in the simplex; see S2 for the other phenotypes).

**Fig 4.**
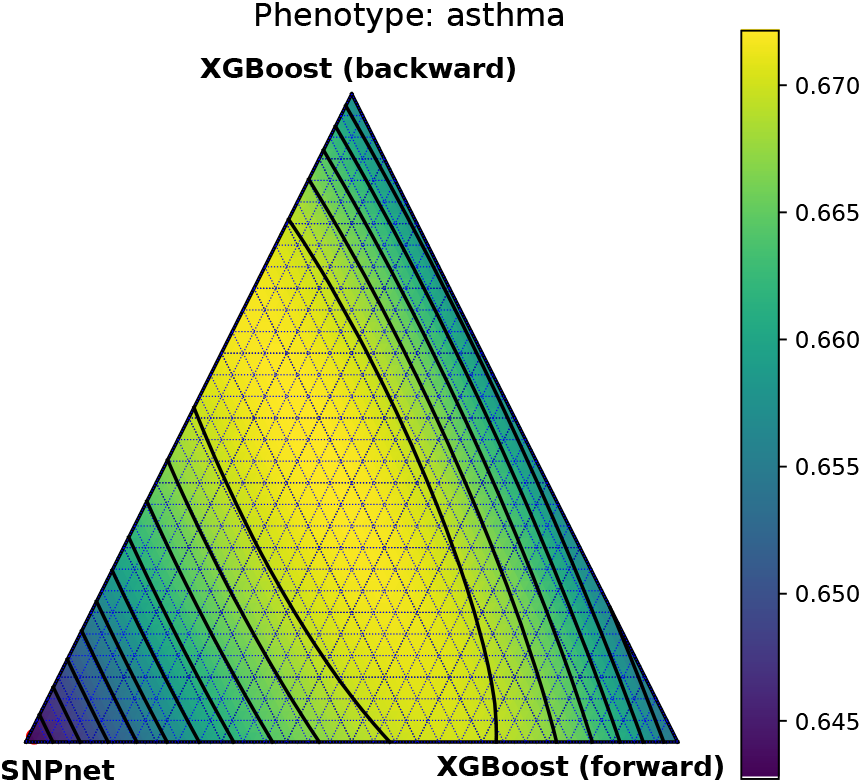
Simplex plot showing the ROC-AUC score obtained by taking weighted sums from three prediction methods: Snpnet (with 50k GWAS-selected SNPs), and XGBoost on 1,000 XGBoost selected SNPs (forward and backward passes). Each vertex of the triangle represents the performance of an individual model.

As already mentioned, we consider two ensembling strategies – a simple ensemble where the sum of predictions is unweighted (corresponding to the simplex center), and stacking, where suitable weights are obtained by maximizing the metric for that phenotype on the validation set. The simple ensemble shows a noticeable improvement in the test metric over the best individual model in all but one phenotype. Stacking always performs as well as or better than the simple ensemble, especially in cases where one method significantly outperforms the rest and the optimal point deviate significantly from the center of the simplex (for example, height in S2).

### 5.3 Analysis of nonlinearity

One can think of nonlinearity in the phenotype-to-phenotype dependence as resulting from three possible kinds of feature interactions: pure gene interactions, gene–covariate interactions, and pure covariate interactions. Since some of the covariates are numerical and so potentially carry complex information, the last group may also include “self-interactions”, in the sense that a phenotype may depend nonlinearly on a particular single covariate. We analyze the three interaction groups one-by-one.

#### Pure gene interactions

This type of interactions should manifest itself in a better performance of the depth-2 XGBoost models compared to the depth-1 models on data not involving additional covariates, i. e. in our ‘Sex-only’ group of results. However, as wee see in Figure 3, the differences between these two models are insignificant in this group for all the considered phenotypes. It is important to note that depth-2 XGBoost model is well suited only for pairwise interactions and not for higher-order ones. Therefore it remains possible that the gene interactions for these phenotypes are high-order.

#### Gene–covariate interactions

The presence of these interactions can be tested by restricting the XGBoost trees to contain either only SNP features or only covariate features (since in this case the model becomes a sum of two terms – one depending on the SNPs and another on the covariates). In Table 1 we compare models trained with or without this restriction. We see that there is almost no difference between such models (and even if present, it is in favor of the restricted model, probably due to an implicit regularization effect).

**Table 1.** Performance of depth-2 XGBoost prediction models with or without restricting gene-covariate interactions

| Phenotype | Metric | w/o restriction | with restriction |
| --- | --- | --- | --- |
| Height | $r^2$ | 0.666 | 0.666 |
| eBMD | $r^2$ | 0.264 | 0.265 |
| Hypothyroidism | AUC | 0.799 | 0.799 |
| Asthma | AUC | 0.656 | 0.659 |
| Psoriasis | AUC | 0.725 | 0.725 |

#### Pure covariate interactions

The presence of these interactions can be tested by comparing XGBoost models of depth 1 and 2 on genotype data with covariates. In this case, we do observe a small increase of accuracy in depth-2 models for all phenotypes except height (see Figure 3).

To further confirm the nonlinear effects of covariates, we compare Snpnet and XGBoost models fitted on the exact same dataset. Table 2 shows performance of these models for common SNP sets selected either by XGBoost or GWAS. While for small number of covariates Snpnet generally outperforms or is on par with XGBoost, for 20+ covariates XGBoost consistently outperforms SNPnet, especially for hypothyroidism and asthma.

**Table 2.**
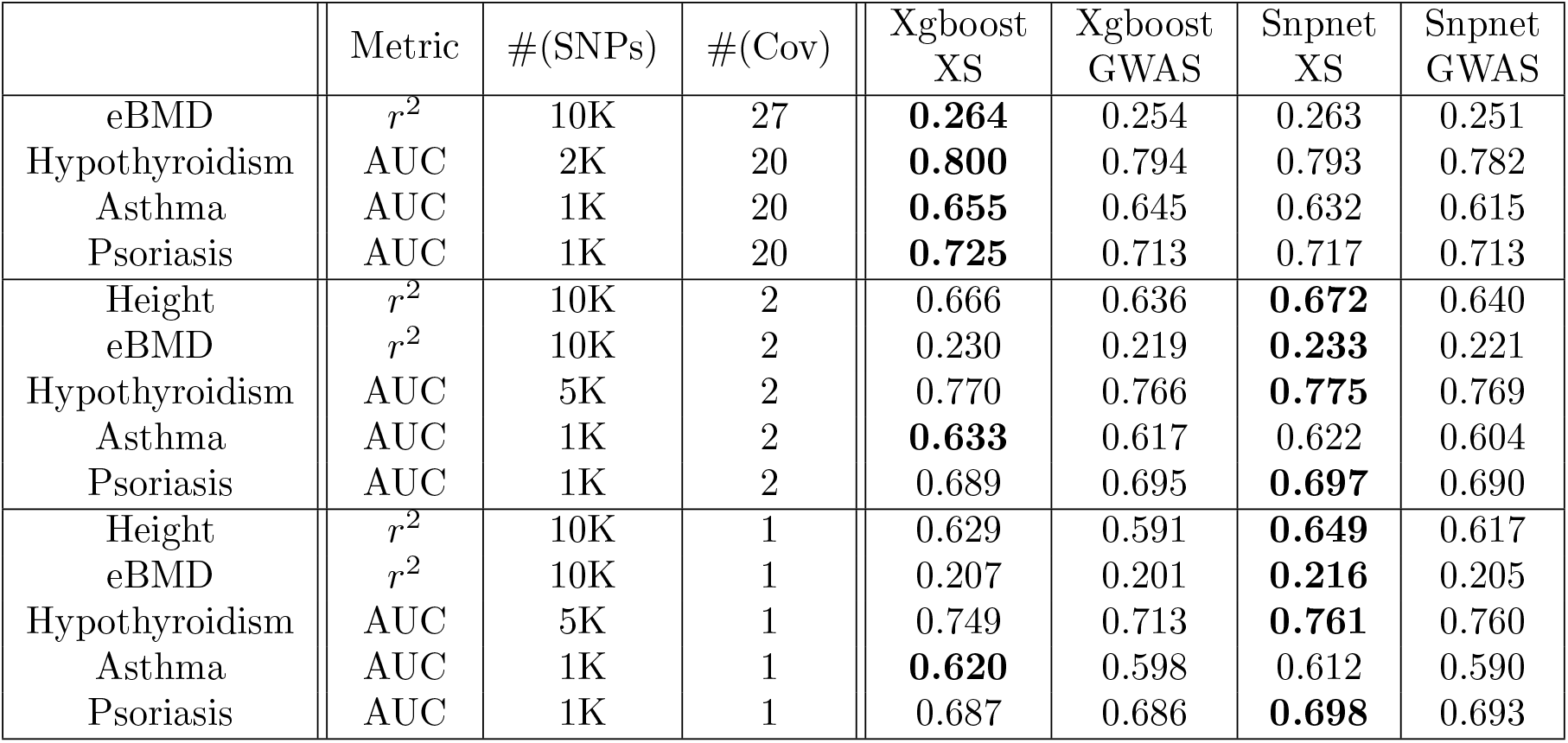
Comparison of GWAS- and XGBoost-based (denoted XS) SNP selections.

### 5.4 Analysis of XGBoost selection

Since SNP selection by XGBoost is our main technical innovation in this paper, we provide now a more in-depth analysis of its properties.

#### 5.4.1 Detection of epistatic interactions

As already discussed, we have not achieved any improvement in prediction accuracy for our five UK Biobank phenotypes by exploiting potential gene-gene and gene-covariate interactions. A natural question is whether this is because such interactions are too weak or because our prediction pipeline is simply not capable of taking advantage of them.

To answer this question, we tested our pipeline on simulated phenotypes. We performed a number of simulations with both linear and epistatic effects and evaluated the detection power of XGBoost feature importance scores. Results are shown in Figure 5. We see that for simulated phenotypes, XGBoost catches most of the epistatic variance and consistently outperforms Lasso. Also, XGBoost is able to detect roughly 55% of epistatic SNPs. Therefore, we expect XGBoost to outperform Lasso on a phenotype with strong enough epistatic effects. Summarizing, we attribute the lack of improvement from using our nonlinear models on the real phenotypes to the lack of nonlinear information in the data rather than to deficiencies of the models.

**Fig 5.**
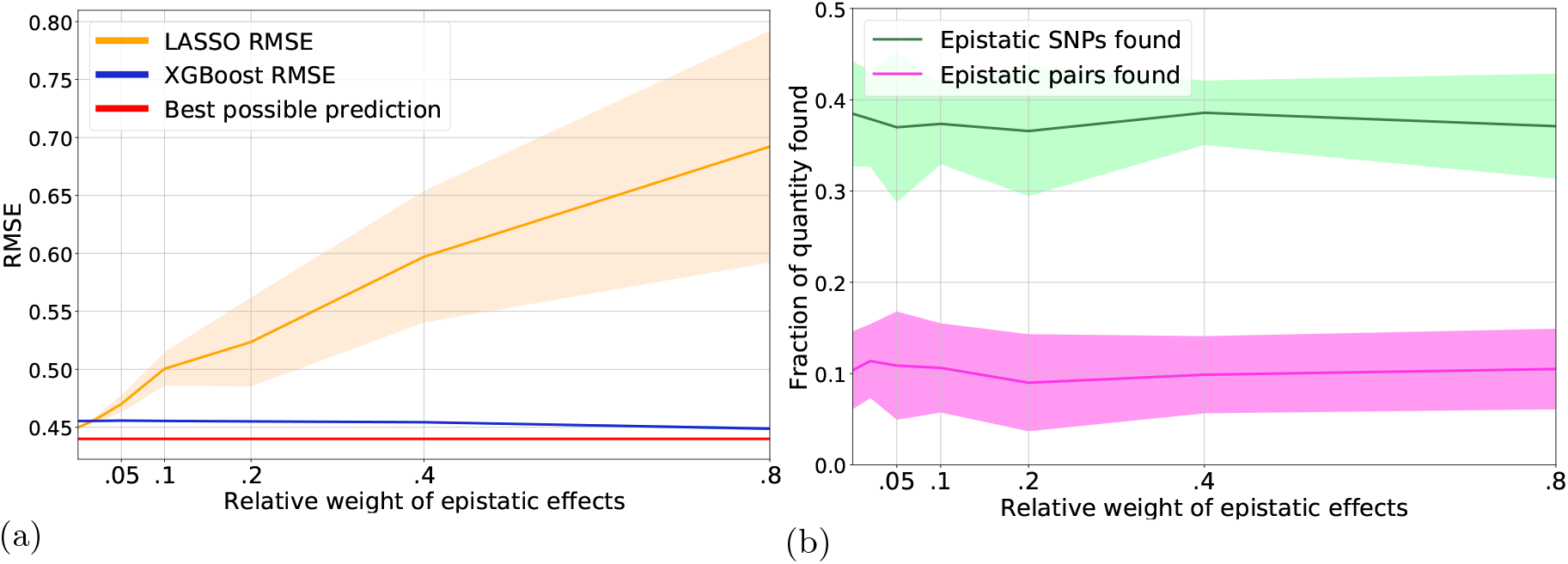
Evaluation of XGBoost prediction and selection performance on simulated data, using 100K samples, 1K SNPs, 100 linearly associated SNPs, 50 epistatic pairs. The weight of random noise is *w*_*ϵ*_ = 0.2, the total weight *w*_*ep*_ of epistatic effects ranges from 0 to 0.8, the total variance of linear effects equals 1 *− w*_*ep*_ *− w*_*ϵ*_, i.e. varies from 0.8 to 0. Shadows of lines have width of two standard deviations. **(a)** Prediction performance of Lasso and XGBoost when varying the relative weight of epistatic effects in the simulated phenotype. **(b)** Fraction of epistatic SNPs and epistatic SNP pairs found by XGBoost. An SNP is considered found when its importance is in the top 300 most important SNPs. A pair is considered found when both SNPs are in the top 300 most important SNPs.

#### 5.4.2 Correlations between selected SNPs

Since XGBoost-based SNP selection takes into account the already chosen SNPs when selecting the next ones, we expect the whole selected set of SNPs to be in an approximate linkage equilibrium. This hypothesis can be tested by comparing correlation between GWAS-selected SNPs with correlations between XGBoost-selected SNPs. The respective correlation matrices for asthma are shown in Figure 6. We see that SNPs selected by XGBoost are significantly less correlated: the mean squared correlation coefficient of the top 1K SNPs is 0.0593 for GWAS and 0.0063 for XGBoost.

**Fig 6.**
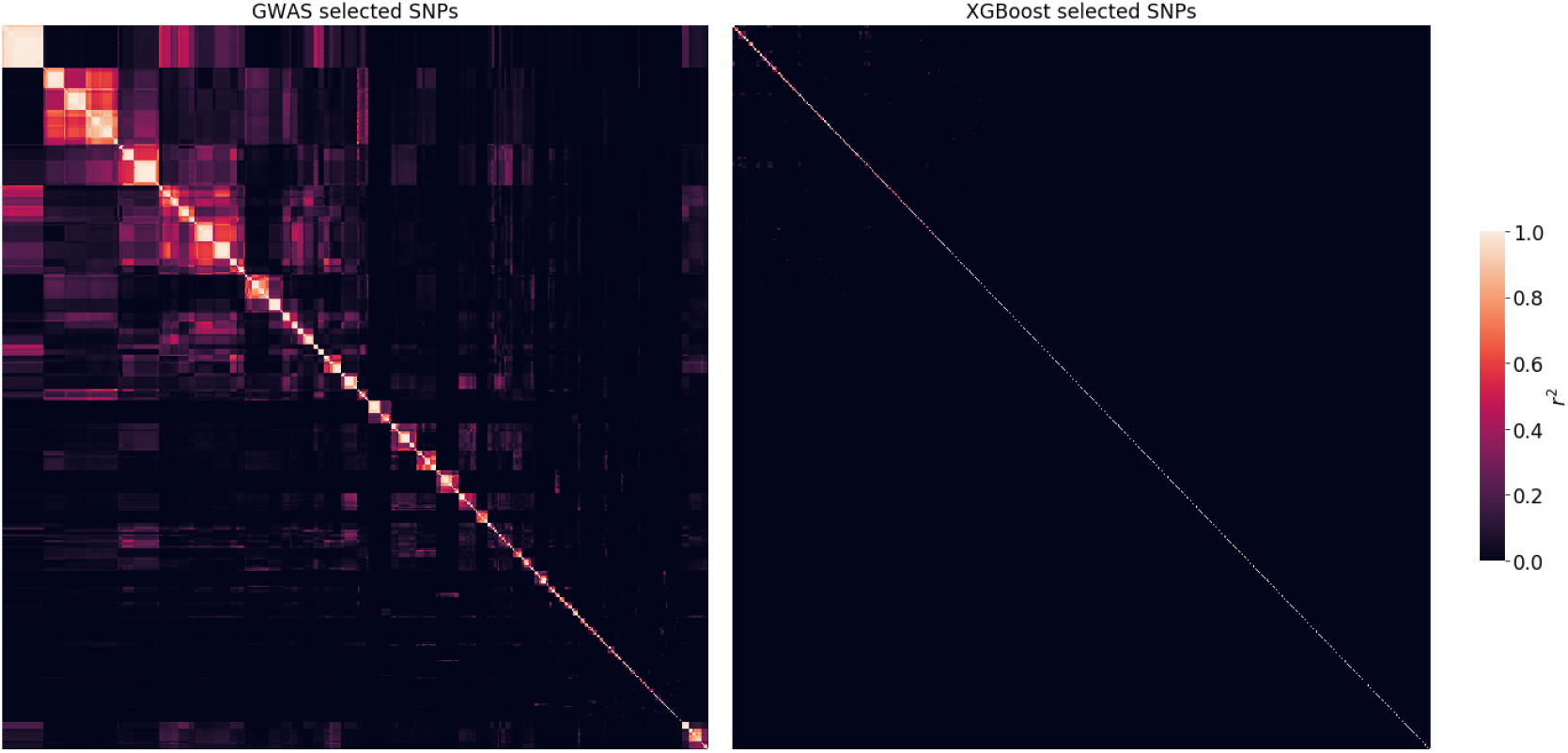
Clustered correlation heatmaps between top 1K SNPs selected by GWAS (**left**) and XGBoost (**right**) for asthma with 20 covariates, sex and PCA. GWAS selects SNPs by lowest p-value, XGBoost selects SNPs by highest importance score. The SNP sets in the left and right subfigure are different; the SNPs are sorted to produce clusters on the heatmaps. The color shows the squared correlation coefficient *r*^2^ between SNPs, estimated by plink 1.9. The average *r*^2^ is 0.0593 for GWAS and 0.0063 for XGBoost.

#### 5.4.3 GWAS vs XGBoost selection for the same number of SNPs

The lower correlation between SNPs selected by XGBoost compared to GWAS suggests that the former selection is more efficient in the sense of producing SNP subsets that are smaller while carrying a comparable amount of relevant information about the phenotype. To test this, we compared performance of these two SNP selection methods at fixed numbers of selected SNPs (Table 2). For each phenotype, the number of SNPs was chosen as the number after which the XGBoost model built on XGBoost-selected SNPs stops improving. In all cases, the dataset of XGBoost-selected SNPs allows to achieve a better accuracy compared to the dataset of GWAS-selected SNPs of the same size. GWAS starts to outperform XGBoost selection only when it selects at least twice the number of SNPs.

Our analysis also shows that XGBoost is better for the final prediction when the number of covariates is big, while Snpnet makes better use of SNP information (Table 2). This finding is consistent with our observation of exploitable nonlinearity present in covariates but not in genotype data.

For a deeper analysis, in Figure 7 we show performance of the four combinations of the two selection methods (GWAS or XGBoost) with the two predictive models (XGBoost or Snpnet), for predicting asthma. The two GWAS-based pipelines show a similar pattern of worse performance on a small set of SNPs and plateauing after roughly 15K SNPs. The other two pipelines work much better on small sets of SNPs but saturate quicker, roughly at 5-7K SNPs. In agreement with results in Figure 3, the XGBoost prediction models are generally better than Snpnet for this phenotype (asthma) at a large number of covariates. This is, of course, not always the case for other phenotypes.

**Fig 7.**
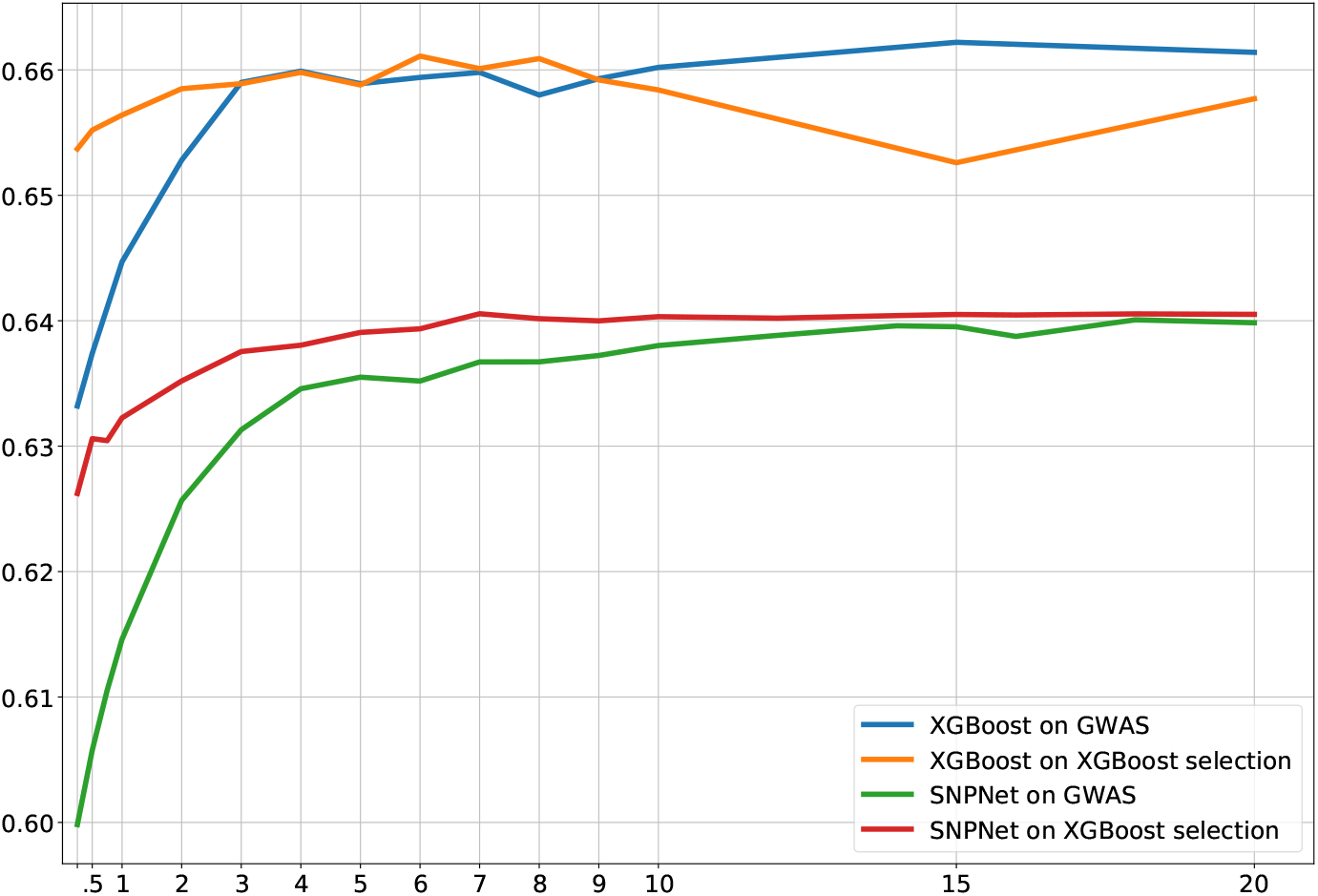
Dependence of the final model accuracy on the number of SNPs for four different combinations of selection and prediction methods for asthma. Both XGBoost selection and XGBoost prediction used depth-2 models. The features include 20 covariates, age, sex, and 10 principal components.

#### 5.4.4 Effects of SNP pre-ordering

In contrast to GWAS selection, XGBoost selection depends on the order in which the SNPs are processed. We checked the importance of this order by running selection on the eBMD phenotype with the natural forward or backward genome order as well as with randomly shuffled windows. On the validation data set, we obtained *r*^2^ = 0.2771 for the forward pass, *r*^2^ = 0.2757 for the backward pass, and *r*^2^ = 0.2716 for the shuffled dataset. The forward+backward ensemble has *r*^2^ = 0.2807, and adding shuffled predictions negligibly improves the results (to *r*^2^ = 0.2809). This shows that the SNP order and local effects do contribute to the performance of XGBoost selection, but their influence is fairly limited. Nevertheless, the models resulting from different orderings are sufficiently diverse to allow some accuracy gain to be achieved by their ensembling.

## 6 Conclusion

### Nonlinearity

Our results for five UK Biobank phenotypes show that gene-gene and gene-covariate interactions in the data are not strong enough to be exploitable by state-of-the-art nonlinear predictive models so as to allow them to outperform best linear models. This conclusion is based on our comparison of various linear and nonlinear models, with and without cross terms. This conclusion is also confirmed by our study of simulated phenotypes with epistatic interactions on the same genotype data: for such phenotypes, prediction accuracy is consistently improved by using nonlinear models. At the same time, we also observe this improvement for real phenotypes if sufficiently many covariates are included as features for predictions. This shows that the covariates contain more useful nonlinear information than SNP-covariate or pure SNP associations.

### SNP selection

We have proposed a new approach to SNP selection, using a sequential construction of XGBoost models on a series of SNP windows and a simultaneous SNP importance estimation. Compared to the conventional GWAS-based SNP selection, our method creates a more informative and less correlated set of SNPs. For small sets of selected SNPs, models built using our selection pipeline consistently outperform those built using the standard GWAS pipeline. Another essential aspect of our approach is its dependence on the order of features during selection: forward and backward passes lead to different selected sets and hence models; we have shown that these models can be sufficiently diverse for their ensembling to yield even better models.

### Most accurate models

For all phenotypes, the most accurate models are obtained by ensembling indepedently trained models (Snpnet, XGBoost). We observe ensembles to be consistently more accurate than individual models, even for those phenotypes (height, eBMD, asthma) where one of the methods significantly outperforms the others. On the whole, ensembling of the state-of-the-art linear (Snpnet) as well as nonlinear (XGBoost depth-2) models seems to boost the overall accuracy by fully retaining the advantages of the constituent models.

Regarding individual (non-ensembled) models, we found that for one of the considered phenotypes (asthma), our nonlinear XGBoost depth-2 model significantly outperformed the state-of-the-art linear model (Snpnet).

## Data Availability

Data can be accessed through the UK Biobank.

http://www.ukbiobank.ac.uk/

## Acknowledgments

Most computations performed in this project were done on the Zhores cluster [23], and we thank the CDISE HPC team for their assistance. This research has been conducted using the UK Biobank Resource under Application Number ‘43661’. Elena Nabieva was supported by the Russian Science Foundation grant 21-74-20160. Dmitry Yarotsky was supported by Russian Science Foundation, grant 21-11-00373. The funders had no role in study design, data collection and analysis, decision to publish, or preparation of the manuscript.

## Supporting information

### S1 Estimation of standard errors

We estimate the standard error of accuracy metrics *r*^2^ and ROC AUC of our predictive models using two approaches that produce similar results. The first approach is applicable to both metrics and is based on subsampling and a natural scaling assumption. The second approach is only applicable to ROC AUC as it uses the particular form of this statistic.

#### S1.1 A subsampling-based estimate

Let *β* be one of the accuracy metrics (i.e., *r*^2^ or ROC AUC). Let *β*_*∗*_ be its true population value, and let *β*_*N*_ be its estimate obtained using a random *N*-element sample. The estimate *β*_*N*_ is a random variable converging to the deterministic value *β* _*∗*_ in the limit *N*→ ∞. We will assume that the estimate *β*_*N*_ is unbiased up to 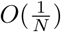:

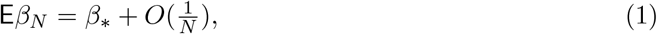

and its variance scales as 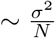 with some *σ*^2^ *>* 0:

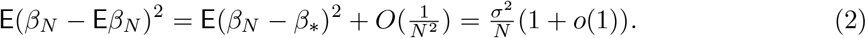

Under these assumptions, for large *N*, we can take the standard error for *β*_*N*_ to be

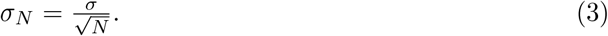

It remains to estimate *σ*^2^. This can be done in the usual way, by considering *M* independent samples *D*_1_, … *D*_*M*_ of size *Ñ*. Denote the respective values of the statistic *β* by *β* _*Ñ*, 1_, … *β* _*Ñ, M*_. Using the standard unbiased estimate of the variance and Eq. (2), and assuming that both *Ñ, M* are large, we can write

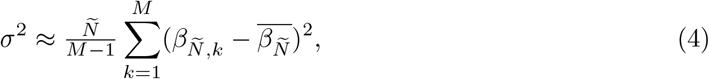

where 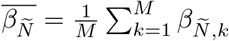. In practice, we form the samples *D*_1_, … *D*_*M*_ by randomly dividing the size-*N* sample into *M* equal parts, so that, in particular, *Ñ* = *N/M*. Combining Eq. (3) with Eq. (4), we conclude that we can estimate the standard error for a size-*N* sample by

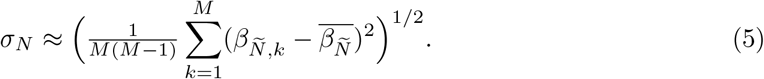

Though this formula depends on *M*, the resulting estimate *σ*_*N*_ should be approximately independent of *M*. In Figure S1 we plot *σ*_*N*_ computed for all our phenotypes at various values of *M*, and indeed observe an approximate independence of *M*, confirming our methodology.

**Fig S1.**
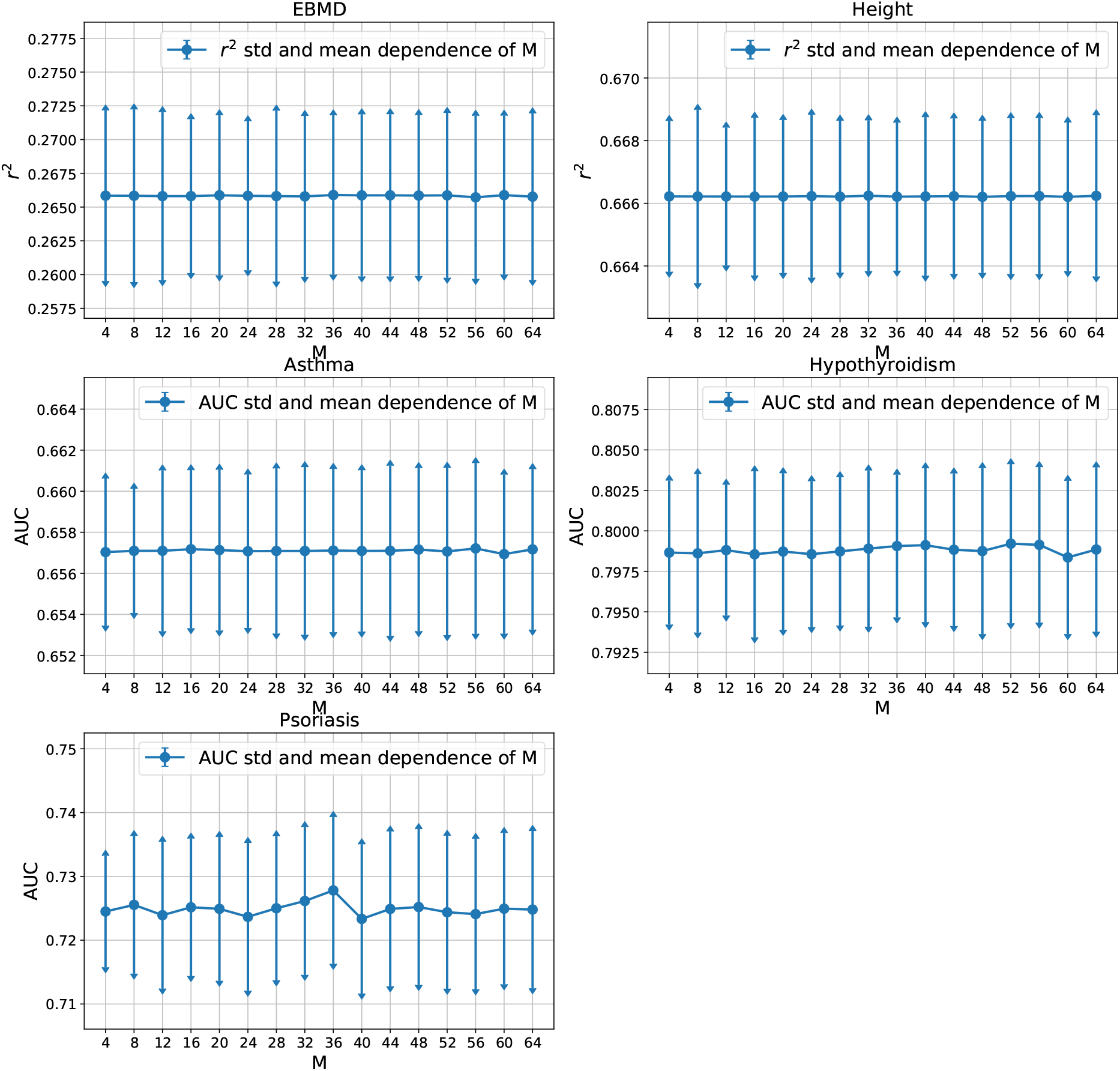
Estimates of the standard deviations *σ*_*N*_ of accuracy characteristics (*r*^2^, ROC AUC) for the 5 phenotypes, as functions of the parameter *M* (see Eq. (5)).

**Table S1.** Standard errors for the accuracy metrics. The columns *σ*_*AUC*_ and *σ*_*N*_ correspond to the two different estimation methods.

| | Metric | $\sigma_{AUC}$ (Eq. (7)) | $\sigma_N$ (Eq. (5)) |
| --- | --- | --- | --- |
| Height | $r^2$ | - | 0.00239 |
| eBMD | $r^2$ | - | 0.00623 |
| Asthma | AUC | 0.00393 | 0.00422 |
| Hypothyroidism | AUC | 0.00474 | 0.00490 |
| Psoriasis | AUC | 0.01140 | 0.01226 |

#### S1.2 Standard error for ROC AUC

Suppose that a “soft” binary classifier produces a numerical value *x*_*k*_ ∈ ℝ for the *k*’th sample of class 0, and a value *y*_*l*_ ∈ ℝ for the *l*’th sample of class 1. The ROC AUC of such a classifier is defined by

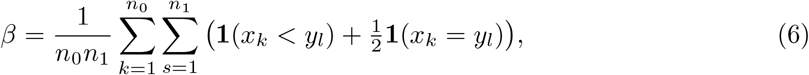

where *n*_0_, *n*_1_ are the numbers of samples in the classes 0 and 1.

Assuming that the samples *x*_*k*_ and *y*_*l*_ are randomly drawn from populations with a continuous distribution function, the variance of this classifier is (see [33, 34])

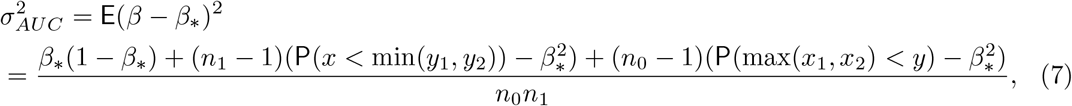

where *β*_*∗*_ = E*β* is the population mean.

Assuming that *n*_0_ and *n*_1_ are large, the leading term of 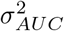 is

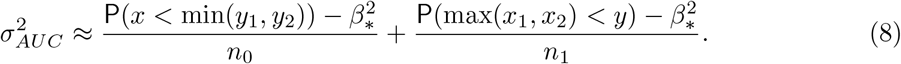

In particular, if *x* and *y* have the same distribution, then 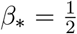 and 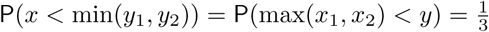, so that

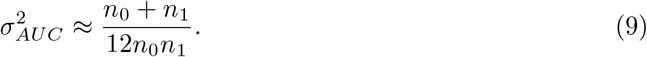

In the general case, when *x* and *y* have different distributions, we can estimate the probabilities P(*x <* min(*y*_1_, *y*_2_)) and P(max(*x*_1_, *x*_2_) *< y*) by sampling. Using additionally the estimated value *β*_*∗*_ of ROC AUC, we can then obtain an estimate for 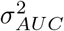.

In Table S1 we provide the standard errors estimated for our phenotypes by the two methods described in this and the previous section. In the case of ROC AUC the results are quite close, confirming the validity of our estimates.

### S2 Simulated phenotypes: simulation details

A simulated phenotype is modelled as a weighted linear combination of linear effects **y**_*l*_, pairwise epistatic effects **y**_*ep*_ and random noise **y**_*ϵ*_:

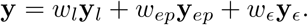

Here, **y, y**_*l*_, **y**_*ep*_, **y**_*ϵ*_ are vectors with *N* components corresponding to different simulated samples. The weights *w*_*l*_, *w*_*ep*_, *w*_*ϵ*_ are sample-independent and sum to 1. The values of the weights depend on the simulation run. For simulation experiments that are presented in this paper we set *w*_*ϵ*_ = 0.2 and vary *w*_*ep*_ from 0 to 0.8.

**Table S2.** Accuracy of XGBoost and SNPnet models constructed for the 5 UK Biobank phenotypes. The SNPs were selected using XGBoost selection for the XGBoost models, and GWAS for SNPnet. The column #SNP shows the number of selected SNP’s for all methods except SNPnet with unlimited SNPs, for which this number is indicated separately.

|  | Metric | #SNP | Covariates | XGBoost<br>depth 1 | Xgboost<br>depth 2 | SNPnet<br>lim. SNPs | SNPnet<br>unlim. SNPs |
| --- | --- | --- | --- | --- | --- | --- | --- |
| eBMD | $r^2$ | 10K | 27 + PCA | 0.258 | 0.264 | 0.251 | 0.280 (50K) |
| Hypothyroidism | AUC | 2K | 20 + PCA | 0.793 | 0.800 | 0.782 | 0.800 (20K) |
| Asthma | AUC | 1K | 20 + PCA | 0.651 | 0.655 | 0.615 | 0.643 (50K) |
| Psoriasis | AUC | 5K | 20 + PCA | 0.720 | 0.725 | 0.713 | 0.726 (10K) |
| Height | $r^2$ | 10K | 2 + PCA | 0.666 | 0.666 | 0.640 | 0.686 (50K) |
| eBMD | $r^2$ | 10K | 2 + PCA | 0.223 | 0.230 | 0.214 | 0.249 (50K) |
| Hypothyroidism | AUC | 5K | 2 + PCA | 0.770 | 0.769 | 0.769 | 0.775 (50K) |
| Asthma | AUC | 5K | 2 + PCA | 0.633 | 0.633 | 0.627 | 0.635 (50K) |
| Psoriasis | AUC | 5K | 2 + PCA | 0.703 | 0.704 | 0.706 | 0.709 (10K) |
| Height | $r^2$ | 10K | 1 | 0.631 | 0.629 | 0.617 | 0.663 (50K) |
| eBMD | $r^2$ | 10K | 1 | 0.208 | 0.207 | 0.205 | 0.233 (50K) |
| Hypothyroidism | AUC | 5K | 1 | 0.751 | 0.749 | 0.742 | 0.767 (20K) |
| Asthma | AUC | 5K | 1 | 0.621 | 0.620 | 0.617 | 0.626 (50K) |
| Psoriasis | AUC | 5K | 1 | 0.700 | 0.705 | 0.707 | 0.711 (20K) |

**Table S3.**
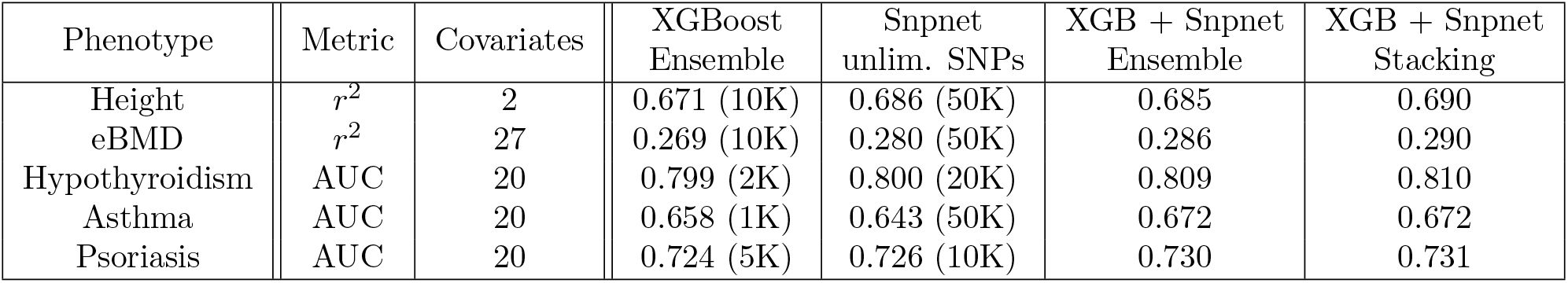
Comparison of best prediction methods. The values in parentheses show the number of pre-selected SNPs in the individual models; these numbers were chosen as providing the best performance. The XGBoost Ensembles consist of two XGBoost models trained on two different sets of SNPs, selected by either forward of backward pass over the genome. The XGB+Snpnet Ensemble and Stacking refer, respectively, to the unweighted or weighted average of the XGBoost forward, XGBoost backward and Snpnet.

**Fig S2.**
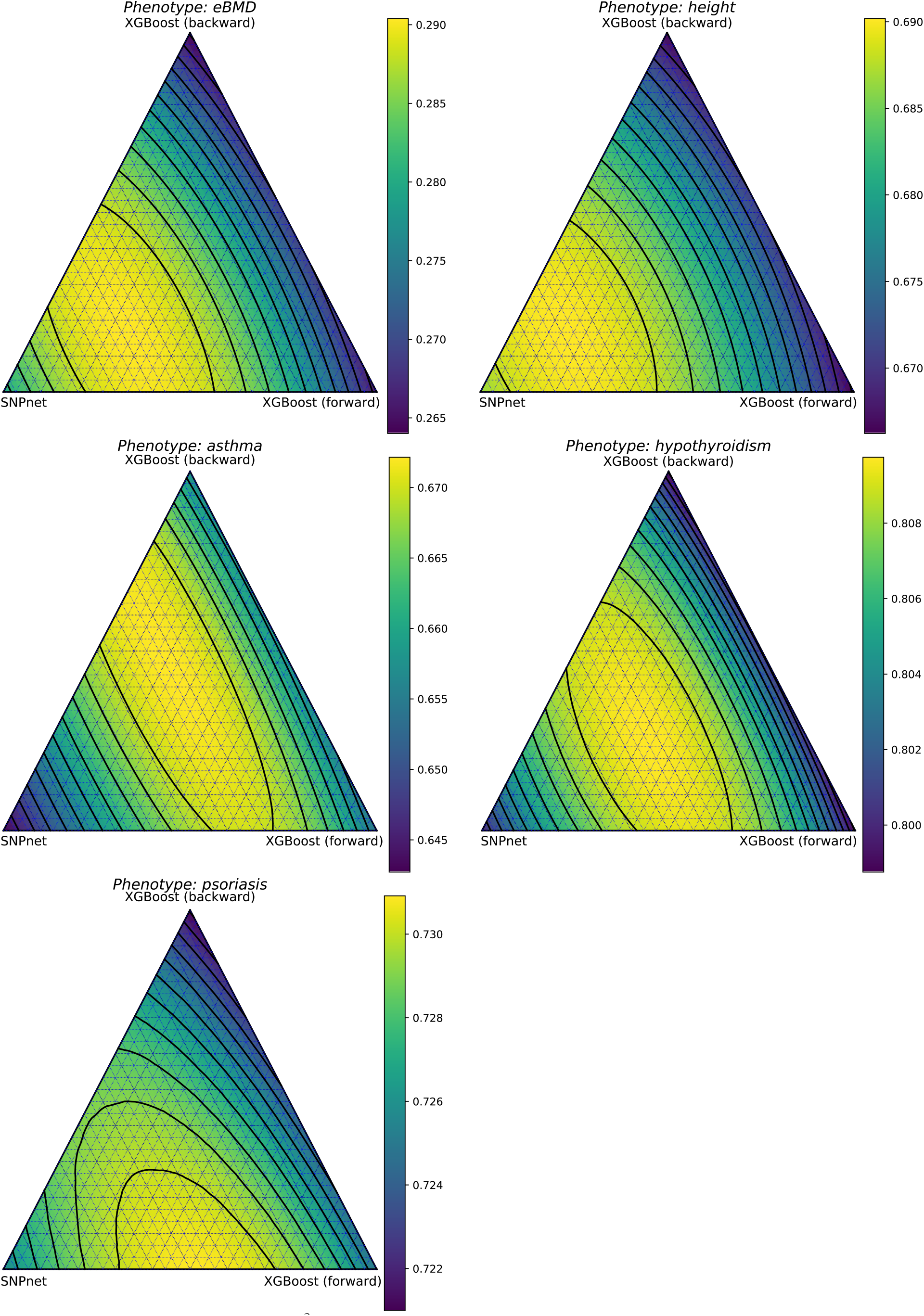
Visualization of metrics (*r*^2^, ROC AUC) for different weighted ensembles of SNPnet, XGBoost forward and XGBoost backward.

Linear effects **y**_*l*_ are generated by **y**_*l*_ = *X***f**_*l*_, where *X* is the *N* × *M* genotype matrix, with *M* the number of SNPs and **f**_*l*_ the *M* -dimensional column vector of individual SNP effects. In all our experiments *M* = 1000 and *N* = 100000. Importantly, **f**_*l*_ has only 0.1*M* = 100 non-zero entries which are distributed normally with zero mean and unit variance. This is done to mimic the sparsity of real-world genotype-phenotype interaction. Linear effects are scaled inversely by MAF (minor allele frequency) of the corresponding SNP to reflect the fact that rarer SNPs typically have larger effect sizes [35]. Non-zero entries of **f**_*l*_ are clipped to the interval [− 5, 5].

To generate pairwise epistatic effects, we randomly select 25 pairs of SNPs from the SNPs already associated linearly with the phenotype, and the same number from the SNPs not associated with the phenotype. For each pair *i* = (*i*_1_, *i*_2_) of SNPs we generate a random epistatic effect size *f*_*ep,i*_ ∼ 𝒩 (0, 1) and randomly select a pair of SNP minor allele counts *c*_1_, *c*_2_, i.e. one value from the set {0, 1, 2} for each SNP in the pair. Then, the epistatic effect for pair *i* and sample *j* is calculated by

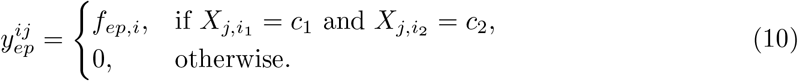

Here, 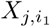 and 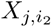 are the minor allele counts of the first and second SNP in the *i*’th pair, respectively. The total value of epistatic effects for sample *j* is

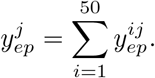

In contrast to the linear effects, the epistatic effects are not scaled inversely with MAF.

**Table S4.**
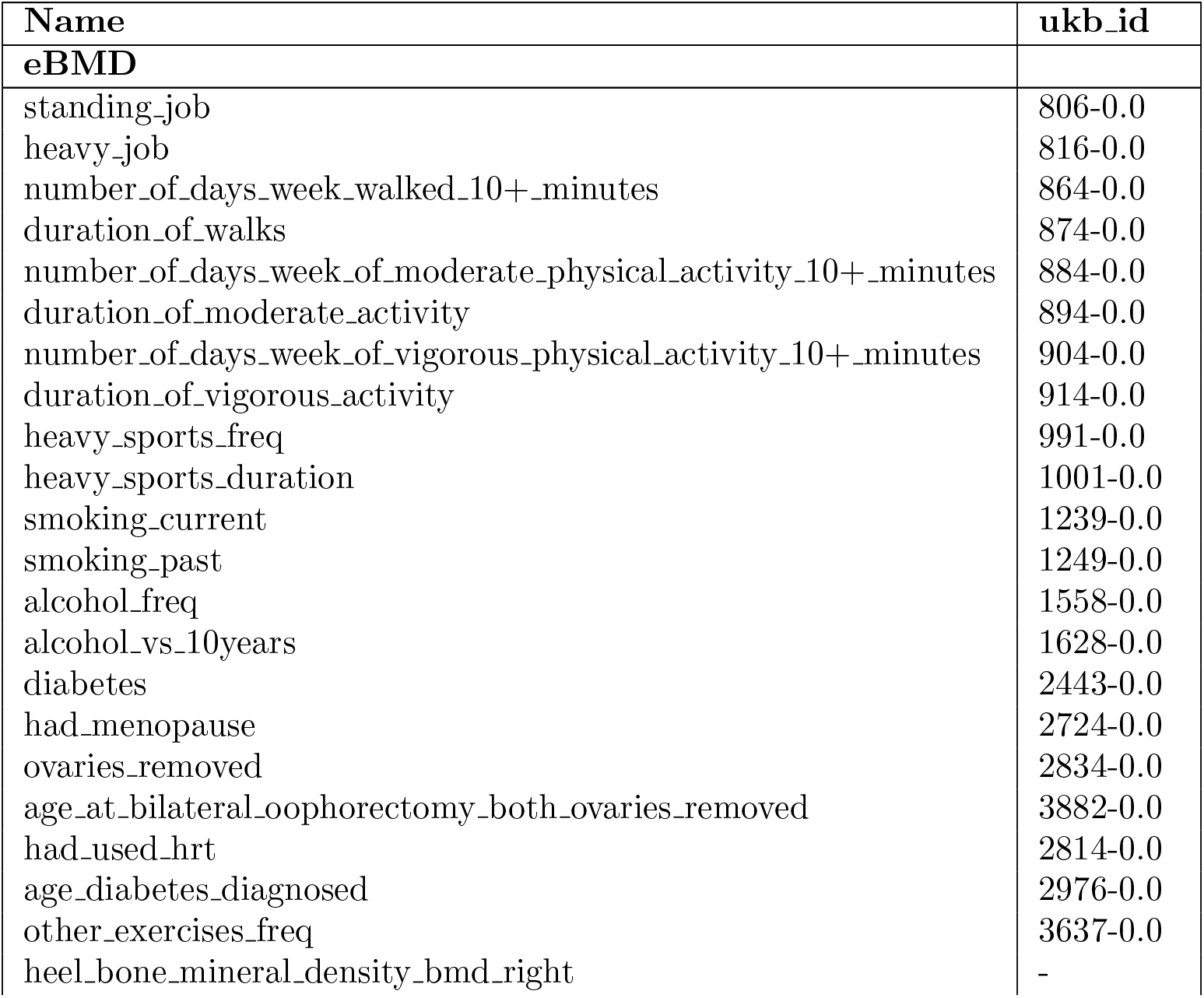
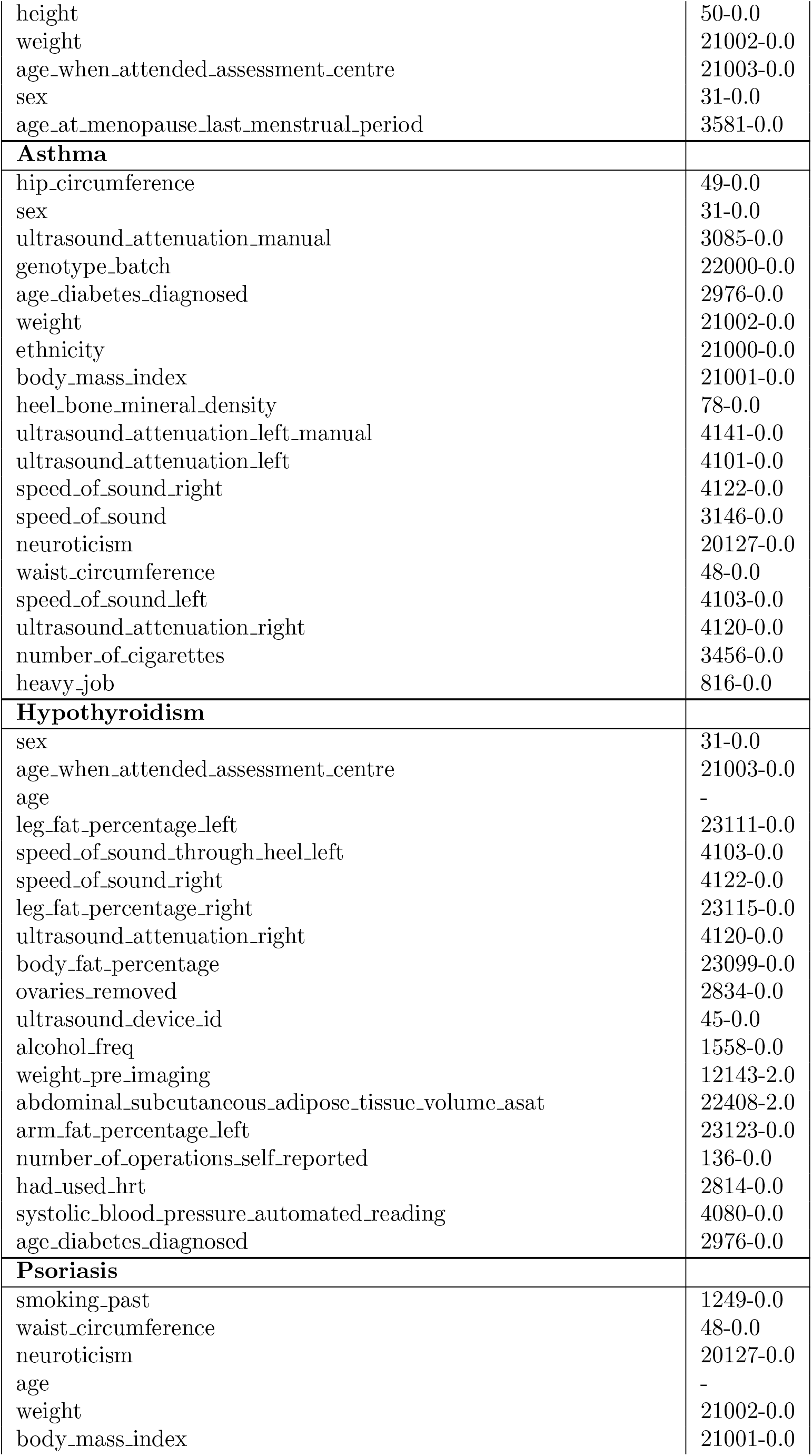
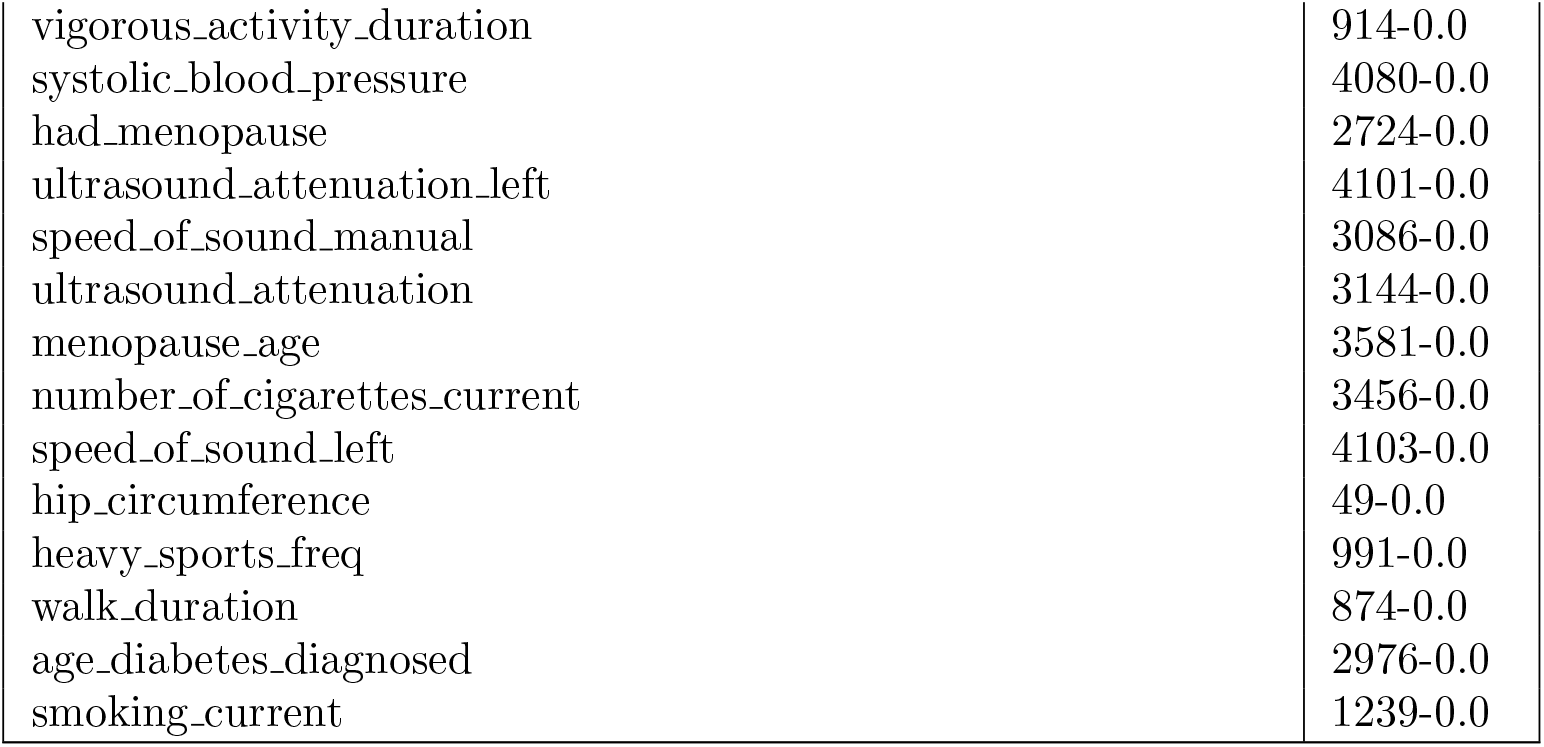
Names and UK Biobank field IDs of additional covariates used in our models for each phenotype.

